# Tolerability of Long-acting Diquafosol Ophthalmic Solution as per Tear Film and Meibomian Glands Findings

**DOI:** 10.1101/2024.05.24.24307866

**Authors:** Reiko Arita, Shima Fukuoka, Minako Kaido

**Affiliations:** Itoh Clinic, Saitama, Japan; Lid and Meibomian Gland Working Group, Saitama, Japan; Omiya Hamada Eye Clinic, Saitama, Japan; Wada Eye Clinic, Chiba, Japan

## Abstract

Long-acting diquafosol (DQS) ophthalmic solution (DQS-LX) has significant advantages regarding patient adherence owing to the reduced frequency of required eye drops; however, some patients prefer conventional DQS over DQS-LX. Herein, to clarify the characteristics of patients according to their preference for ophthalmic solutions, dry eye (DE) and meibomian gland (MG) findings were retrospectively investigated. This study enrolled 341 patients with DE (mean age, 62.1 ± 11.7 years) treated at the Itoh Clinic between November 8, 2022, and July 31, 2023, who switched from DQS to DQS-LX. Patients were divided into two groups: those who continued DQS-LX administration (DQS-LX group) and those who wished to revert to conventional DQS (DQS group). Data regarding subjective symptoms assessed using the Standard Patient Evaluation of Eye Dryness (SPEED) questionnaire, tear film breakup time (BUT), tear meniscus height (TMH), corneal and conjunctival fluorescein staining (CFS), conjunctival hyperemia/papilla, meiboscore, plugging, vascularity, meibum grade, and Schirmer’s score at the time of DQS-LX switch were evaluated. Of the 341 patients, 31 (9.1%) wished to revert to conventional DQS. In total, 32 eyes of 16 patients in the DQS group and 64 eyes of 32 patients in the DQS-LX group—for whom complete data were available—were included in the analysis. Compared with the DQS group, the DQS-LX group had significantly higher SPEED scores, shorter BUTs, lower TMHs, greater CFS findings, larger meibum grades (all *P* < 0.001), lower Schirmer scores (*P* = 0.008), and more pluggings (*P* = 0.001) than the DQS group. More allergic conjunctivitis-related complications were observed in the DQS group (*P* = 0.034). In conclusion, patients with low tear film volume and DE complicated by moderate or severe MG dysfunction preferred DQS-LX, while those with allergic findings preferred conventional DQS.

## Introduction

Tear film instability, in which an imbalance in the ocular surface tear film deteriorates its stability and regularity, is an important cause of dry eye (DE). The P2Y_2_ receptor agonist diquafosol tetrasodium (DQS) was developed as an eye drop to improve tear film stability by stimulating tear and mucin secretion [1, 2]. The introduction of this ophthalmic solution has led to significant changes in treatment strategies for patients with DE. Many reports have shown the efficacy of DQS ophthalmic solutions in improving corneal staining, stabilizing tear fluid film, and relieving DE symptoms [3–6]. As P2Y_2_ receptors are also present in meibomian glands, a DQS-induced increase in the lipid layer has been reported in several studies [7–12]. DQS comprehensively targets the aqueous, mucin, and lipid layers. However, DQS ophthalmic solutions must be applied six times per day to keep the ocular surface moist, which reduces patient compliance. Uchino et al. reported that only 10.2% of participants applied eye drops at the frequency described in the package insert [13].

Recently, a new long-acting formulation of DQS ophthalmic solution (DQS-LX) was developed [14] through the addition of polyvinylpyrrolidone (PVP), which reduced the required frequency of eye drops to three times a day. While several patients who were prescribed DQS switched to DQS-LX, some preferred DQS [15, 16]. Previous studies have reported changes in subjective symptoms, fluorescein tear film breakup time (FBUT), and fluorescein staining findings after switching from DQS to DQS-LX [15, 16]. However, no comprehensive studies have investigated ocular surface parameters, including meibomian gland-related parameters and allergic conjunctivitis-related complications.

Herein, we compared ocular surface parameters, including the meibomian gland findings, related to the parameters and condition of the tarsal conjunctiva, to identify differences between patients who were on conventional DQS and switched to DQS-LX, those who were satisfied with DQS-LX and wished to continue DQS-LX, and those who wished to revert to DQS.

## Materials and Methods

This retrospective cohort study was approved by the Institutional Review Board of the Itoh Clinic and adhered to the tenets of the Declaration of Helsinki (Registration ID: IRIN2023-0909). Informed consent was obtained from all the participants. This study was registered with the University Hospital Medical Information Network (Registration ID: UMIN000054378). There was no contact with patients or legal guardians as all data was obtained by study investigators through patient identification numbers on the electronic health system and subsequently fully anonymized to ensure patient confidentiality.

### Participants

This study included 341 DE patients (62.1 ± 11.7 years) treated at the Itoh Clinic between November 8, 2022, and July 31, 2023 who switched from DQS to DQS-LX. Data of those who continued DQS-LX for at least 1 month (DQS-LX group) and those who wished to revert to conventional DQS within 3 months (DQS group) were retrospectively compared. Patients using eye drops or oral medications other than DQS were included in the current study, and other treatments remained unchanged, except for the switchover from DQS to DQS-LX. Both eyes of patients were included in this study. Patients who had used DQS for less than 3 months, used DQS-LX for less than 1 month, had punctal plugs, wore contact lenses, used anti-glaucoma eye drops, undergone eye surgery within 3 months, and could not provide consent were excluded.

### Clinical Assessment

Clinical assessment items were measured at the time of the DQS-LX switchover. Symptoms were assessed using the Standard Patient Evaluation of Eye Dryness (SPEED) validated questionnaire (scale, 0–28) [17,18]. Tear meniscus height (TMH) was quantitatively measured using an IDRA (SBM Sistemi, Torino, Italy) [17, 18]. Lid margin abnormalities (plugging of the meibomian gland orifices and vascularity of lid margins) [19], FBUT, corneal and conjunctival fluorescein staining (CFS) [20], and meibum grade (0-3) [21] were evaluated using slit-lamp microscopy. FBUT was measured after instilling 1 μl of preservative-free 1% sodium fluorescein into the conjunctival sac using a micropipette. CFS was scored on a scale of 0–9 points as previously described [20]. Conjunctival hyperemia and papillae were observed. Morphological changes in the meibomian glands were assessed based on the meiboscore (0-6) [22] as determined by noninvasive meibography. Tear fluid production was measured using Schirmer’s test without anesthesia [23].

### Statistical Analysis

Data are presented as means ± standard deviation (SD). The Shapiro–Wilk test revealed the non-normal distribution of the data (*P* < 0.05); thus, nonparametric tests were used. The Fisher’s exact test was used to compare categorical variables between the DQS and DQS-LX groups. The Mann–Whitney *U* test was used to compare continuous variables between the two groups. We performed a post-hoc power analysis for the SPEED score, TMH, and FBUT. For the SPEED score, the mean difference between the two groups was 3.8, with a corresponding SD of 6.2; for the TMH, the mean difference was 0.04 with an SD of 0.05; and for the FBUT, the mean difference was 0.49 with an SD of 1.23. These changes were calculated using the data from 96 eyes of 48 patients. The power (1 − β) was > 0.95 at the level of α = 0.05 for the SPEED score, TMH, and FBUT, and the sample size of this study was sufficient. Statistical analyses were performed using JMP Pro version 17 software (SAS, Cary, NC, USA). All statistical tests were two-sided, and a *P*-value < 0.05 was considered statistically significant.

## Results

### Demographics of the Study Population

Of 341 patients, 31 (9.1%) wished to revert to conventional DQS. In total 48 (14.1%) out of 341 patients had adequate records and were eligible for further analyses. The DQS group included 32 eyes of 16 patients (64.1 ± 12.6 years) and the DQS-LX group included 64 eyes of 32 patients (61.1 ± 11.2 years) (Table 1). The concomitant therapies and comorbidities at the time of the switchover to DQS-LX are shown in Tables 1 and 2. Regarding complications, allergic conjunctivitis was significantly more common in the DQS group (*P* = 0.034) (Table 1), and significantly more patients in the DQS group used anti-allergic eye drops (*P* = 0.034) (Table 2). Significantly more patients in the DQS-LX group used 0.1% fluorometholone eye drops (*P* = 0.012) (Table 2). Significantly more patients in the DQS-LX group used azithromycin eye drops (*P* = 0.002) and had a significantly higher history of intense pulsed light treatment (*P* = 0.013) (Table 2).

**Table 1.**
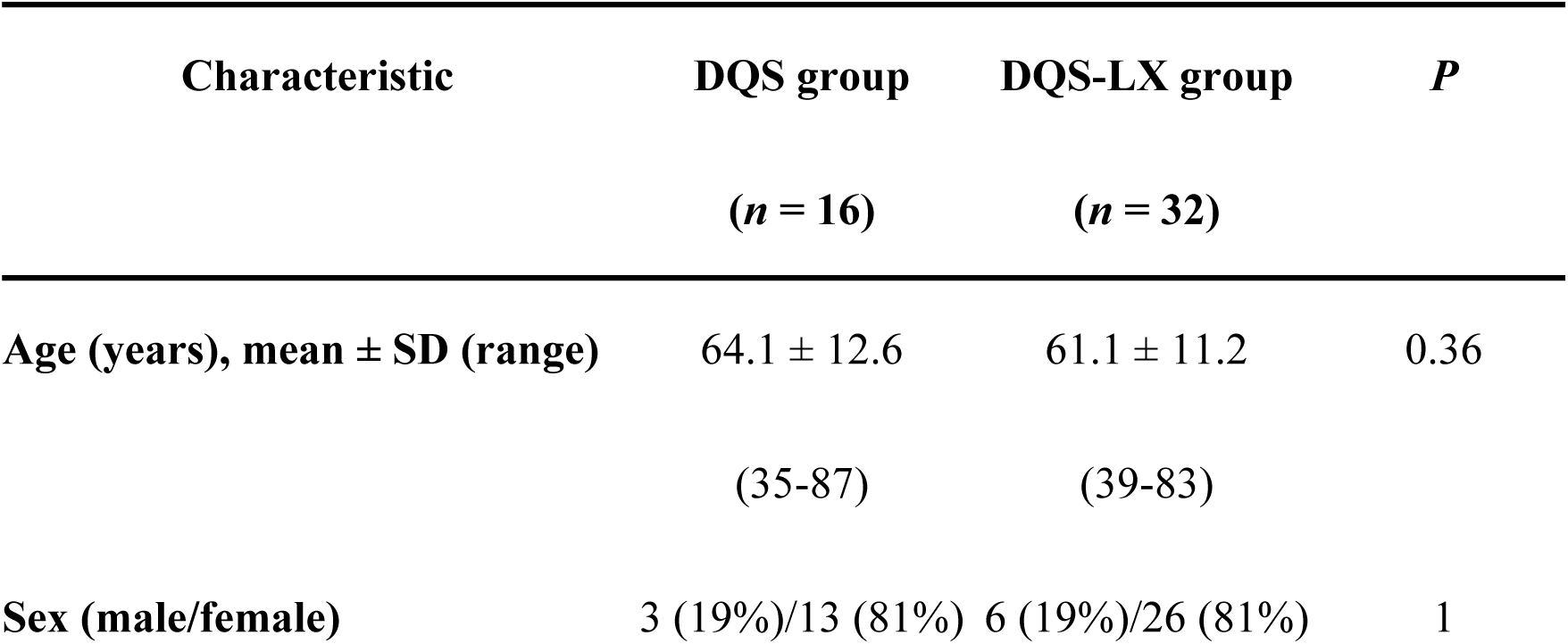
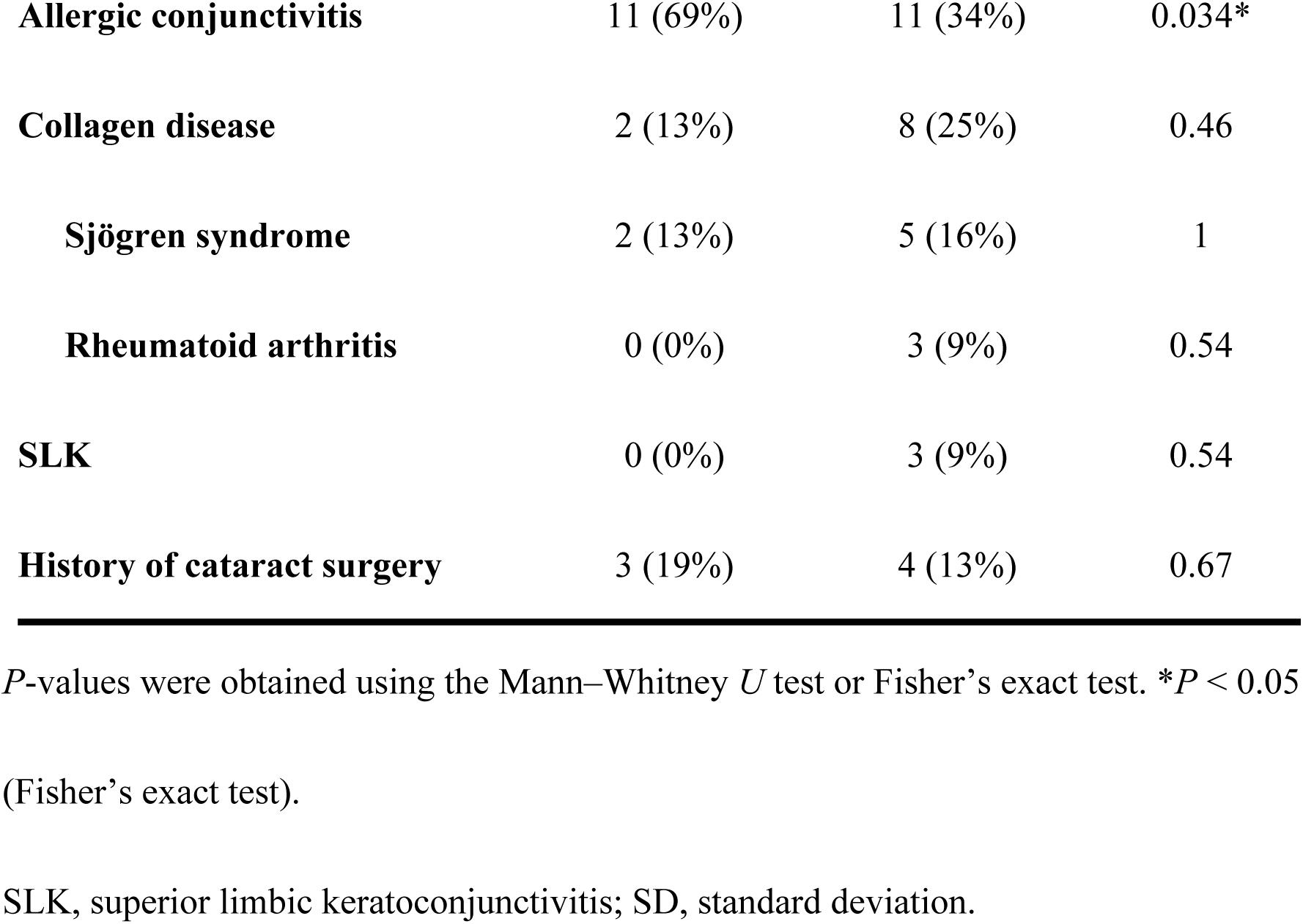
Baseline characteristics of patients in the DQS and the DQS-LX groups.

**Table 2.** Concomitant therapies at the switchover from DQS to DQS-LX in the DQS and DQS-LX groups.

| **Therapy** | **DQS group**<br>**(*n* = 16)** | **DQS-LX group**<br>**(*n* = 32)** | ***P*** |
| --- | --- | --- | --- |
| **Eye drops for dry eye** | 2 (13%) | 10 (31%) | 0.29 |
| **Rebamipide UD** | 2 (13%) | 7 (22%) | 0.70 |
| **Sodium hyaluronate 0.1%** | 0 (0%) | 3 (9%) | 0.54 |
| **Sodium hyaluronate Mini 0.3%** | 0 (0%) | 1 (3%) | 1.0 |
| **Anti-allergic eye drops** | 11 (69%) | 11 (34%) | 0.034* |
| **Epinastine 0.05%** | 1 (6%) | 0 (0%) | 0.33 |
| **Epinastine 0.1%** | 9 (56%) | 10 (31%) | 0.12 |
| **Olopatadine** | 1 (6%) | 1 (3%) | 1.0 |
| **Azithromycin** | 0 (0%) | 14 (44%) | 0.002* |
| **Fluorometholone 0.1%** | 5 (31%) | 23 (72%) | 0.012* |
| **IPL** | 3 (19%) | 19 (59%) | 0.013* |

\* $P < 0.05$ . $P$ -values were obtained using the Fisher's exact test.

Rebamipide UD, rebamipide (Mucosta®) ophthalmic suspension UD2% (unit dose); Sodium hyaluronate 0.1%, purified sodium hyaluronate (Hyalein®) ophthalmic solution 0.1%; Sodium hyaluronate Mini 0.3%, purified sodium hyaluronate single-dose unit (Hyalein® Mini) ophthalmic solution 0.3%; Epinastine 0.05%, epinastine hydrochloride (Alesion®) ophthalmic solution 0.05%; Epinastine 0.1%, epinastine hydrochloride (Alesion® LX) ophthalmic solution 0.1%; Azithromycin, azithromycin hydrate (Azimycin®) ophthalmic solution 1%; Fluorometholone 0.1%, fluorometholone (Flumetholon®) ophthalmic suspension 0.1%; IPL, intense pulsed light therapy.

### Subjective Symptoms and Ocular Surface Parameters at the Time of the Switchover

Subjective symptoms and tear film parameters at the time of the switchover are shown in Table 3. Compared with the DQS group, the DQS-LX group had higher SPEED scores, lower TMHs, shorter FBUTs, greater CFS findings, larger meibum grades (all *P* < 0.001), lower Schirmer scores (*P* = 0.008), and more pluggings (*P* = 0.001). Meiboscores and vascularity did not differ significantly between the two groups (*P* = 0.26 and 0.21, respectively) (Table 3).

**Table 3.** Comparison of subjective symptoms and ocular surface parameters at the switchover from DQS to DQS-LX in the DQS and DQS-LX groups.

| Characteristic | DQS group | DQS-LX group | *P* |
| --- | --- | --- | --- |
| **SPEED (0-28)** | 8.1 ± 4.2 | 16.1 ± 6.9 | <0.001** |
| **TMH (mm)** | 0.24 ± 0.05 | 0.15 ± 0.04 | <0.001** |
| **Plugging (0-3)** | 2.1 ± 0.9 | 2.6 ± 0.7 | 0.001* |
| **Vascularity (0-3)** | 1.3 ± 1.1 | 1.6 ± 1.0 | 0.21 |
| **FBUT (sec)** | 3.3 ± 1.1 | 2.3 ± 1.3 | <0.001** |
| **CFS (0-9)** | 0.3 ± 0.4 | 1.9 ± 2.3 | <0.001** |
| **Presence of conjunctival hyperemia and papillae (n (%))** | 11 (69%) | 11 (34%) | 0.034<sup>†</sup> |
| **Meibum grade (0-3)** | 1.3 ± 0.7 | 2.0 ± 0.6 | <0.001** |
| **Meiboscore (0-6)** | 2.5 ± 1.3 | 2.9 ± 1.2 | 0.26 |
| **Schirmer's test (mm)** | 5.9 ± 4.8 | 3.9 ± 3.6 | 0.008* |

DQS group ( $n = 16$ ) and DQS- LX group ( $n = 32$ ) for SPEED score; DQS group ( $n = 32$ eyes) and DQS- LX group ( $n = 64$ eyes) for the other parameters. Data are presented as means ± SD unless noted otherwise. \* $P < 0.05$ and \*\* $P < 0.001$ (the Mann–Whitney $U$ test). <sup>†</sup> $P < 0.05$ (the Fisher's exact test).
SPEED, Standardized Patient Evaluation of Eye Dryness; TMH, tear meniscus height; FBUT, breakup time of the tear film with fluorescein; CFS, corneal and conjunctival fluorescein staining; SD, standard deviation.

The reasons for reverting to DQS after switching to DQS-LX were stickiness in the eye in the morning (*n* = 13, 3.8%), increased eye discharge (*n* = 12, 3.5%), itchiness after eye drops (*n* = 8, 2.3%), wanting to eye drops use more than three times per day (*n* = 4, 1.2%), and eye irritation (*n* = 3, 0.8%).

## Discussion

The study demonstrated that approximately 90% of patients who were prescribed DQS could tolerate the change to DQS-LX. Furthermore, patients with more severe DE and those with DE complicated by MG dysfunction could tolerate DQS-LX, whereas those with DE complicated by allergic conjunctivitis could not.

The actions of DQS-LX include the following: (i) temporary coating of the ocular surface due to the increased viscosity of the ophthalmic solution caused by PVP, (ii) improved adherence due to the reduced frequency of required eye drops, (iii) mucin and aqueous layer secretion effects of DQS, (iv) an increased lipid layer due to lipid secretion effects, and (v) reduced friction. As DQS and DQS-LX both influence aqueous, mucin, and lipid layers, the efficacy of the eye drops in patients with severe DE may particularly be influenced by the other aforementioned factors. An increase in the lipid layer was observed for DQS in previous reports [7, 8, 11, 24]; however, DQS-LX is considered to facilitate the contact between the liquid layer and the lipid reservoir, increasing the amount of lipids in the tear film. Moreover, the improved adherence may likely lead to better results regarding mucin and water secretion effects and the lipid layer for DQS-LX than for DQS.

Herein, the tolerability for DQS-LX was very good (> 90%). The group with better tolerability to DQS-LX had worse tear films, meibomian gland parameters, and subjective symptoms at the time of the switchover than the group with lesser tolerability to DQS-LX. Patients with moderate-to-severe DE and DE complicated by MG dysfunction tended to tolerate DQS-LX well. We speculated that the increased viscosity of DQS-LX due to the addition of the PVP resulted in a better coating of the ocular surface, reduction in ocular surface irritation, improvement in tear fluid stability, and improvements in corneal and conjunctival epithelial damage. It has also been suggested that DQS-LX may be longer-lasting in patients with DE with concomitant MG dysfunction because of its lipid-increasing effect on the tear film, which decreases friction between the eyelid and cornea, facilitates eyelid opening, and decreases the evaporation of tear fluid.

It has been suggested that patients with DE and allergic conjunctivitis may have difficulty tolerating DQS-LX because PVP, which is added to DQS-LX as a viscosifying agent, increases the residence time of allergens on the ocular surface and exacerbates the symptoms (itching).

Ishikawa et al.[15] reported that 94.4% (51/54) of patients preferred DQS-LX. However, therein[15], only patients with relatively mild disease who used DQS and DQS-LX as a single therapy were included. To represent patients with DE in a real clinical scenario, we included patients who were receiving concomitant DE medications, such as rebamipide or sodium hyaluronate eye drops, as well as patients with a history of intense pulsed light treatment. Although there were differences in disease severity among participants, the results were similar. In a report by Kaido and Arita [16], 84.8% (46/56) of patients had an FBUT of 2.9 ± 1.9 s and a similar DE severity; however, the mean age of the cohort was 74.0 ± 10.4 years, which was older than that in our study cohort. With aging, tear fluid clearance is expected to decrease owing to increased complications of conjunctival chalasis [25] and reduced blinking ability [26]. When these complications occur in older patients, the high viscosity of DQS-LX may result in ineffective diffusion on the ocular surface and excessive retention of ophthalmic fluid in the lower eyelid. The stability of the tear film and improvement of corneal flaws may be affected, making eye drops more difficult to apply and less successful in covering the ocular surface.

The reasons for not tolerating the DQS-LX were as follows: dislike of sticky eyelids upon waking, concern regarding large amounts of eye discharge, itchy eyes, desire for frequent application, and eye irritation. These results are similar to those of previous reports [14, 15]. However, these percentages were similar to the results of Ishikawa et al.[15], but less than those reported by Kaido and Arita.[16] The difference in the proportion of side effects may be related to age differences among the subjects[16]. The Dry Eye Assessment and Management (DREAM) study[27] reported that as age increased, corneal and conjunctival staining findings increased, BUT shortened, symptoms worsened, and tear osmolarity increased; the older age of the previous study’s cohort [16] likely contributed to the higher rate of complaints regarding ocular symptom [28].

The retrospective nature of this study presents a limitation because it examined the tolerability of DQS-LX in patients who were already prescribed DQS. Future prospective studies are required to determine the characteristics of patients who prefer DQS over DQS-LX and vice versa.

## Conclusion

High-viscosity DQS-LX ophthalmic solution is well-tolerated in patients with DE. Patients with moderate or severe DE and MG dysfunction tended to prefer DQS-LX, whereas those with DE and allergic findings preferred conventional DQS.

## Data Availability

All relevant data are within the manuscript and its tables.

